# Immunobridging Analysis of Pemivibart for the Treatment of COVID-19 – A Therapeutic Gap for the Immune Compromised Remains

**DOI:** 10.1101/2025.07.23.25332095

**Authors:** Anna Holmes, Kristin Narayan, Ilker Yalcin, Leijun Hu, Mark A. Wingertzahn

**Author notes:** **Corresponding Author:** Mark A. Wingertzahn, Ph.D.

## Abstract

**Introduction:** SARS-CoV-2 virus inflicts a major ongoing medical toll via acute infection with substantial morbid and mortal outcomes, as well as post-acute sequelae. Multiple SARS-CoV-2 RBD-directed mAbs, which emulate native human immunobiology following infection, have been developed, authorized, and demonstrated substantial efficacy for treatment of COVID-19. However, rapid virus evolution challenges traditional development pathways, as non-susceptible variants can emerge faster than development and regulatory review can be completed. Fortunately, mAb antiviral activity across viral variants can be measured via clinical serum virus neutralizing antibody titers that correlate to demonstrated clinical outcomes from historical mAbs and serve as surrogate biomarkers for efficacy. This analytic immunobridging approach is useful for rapid assessment of novel mAb efficacy, especially for mAbs engineered from a clinically established molecular ancestor. Immunobridging is commonly used in vaccine development and supported the Emergency Use Authorization (EUA) of pemivibart, a mAb targeted to the spike protein of SARS-CoV-2 for prevention of COVID-19 in immunocompromised patients. We therefore applied a similar framework to evaluate pemivibart for the treatment of acute COVID-19, aiming to address the urgent unmet needs of immunocompromised patients who remain vulnerable despite vaccination and antiviral therapies, but failed to secure FDA authorization.

**Methods:** Three complementary methods were used to evaluate immunobridging of pemivibart against 4 dominant variants (JN.1, KP.3.1.1, XEC, and LP.8.1) for the treatment of COVID-19: 1) strict immunobridging of neutralizing antibody titers of pemivibart to its parent molecule adintrevimab, 2) benchmarking comparison of neutralizing antibody titers of pemivibart to other historical mAbs with prior demonstrated efficacy in the treatment of COVID-19, and 3) dose-response analysis of pemivibart to comparator mAbs based on a published meta-analysis.

**Results:** Pemivibart demonstrated strict immunobridging to adintrevimab from 4 to >14 days, depending on variant analyzed. Neutralizing titers of pemivibart across variants were 4-12-fold higher than titers demonstrated for sotrovimab and less than titers for other historical IV-administered mAbs throughout a 14-day analysis period. The dose-response analysis predicted pemivibart to have equivalent efficacy to all other comparator mAbs.

**Conclusion:** Following a similar approach to prevention, immunobridging was demonstrated for pemivibart for the treatment of COVID-19 using multiple complementary methods and suggesting substantial antiviral activity that would accelerate and amplify the native human antibody response. This development methodology, which draws on quantitative virology, biological mechanism, and the U.S. regulatory response to our initial approach provides a road map for accelerating novel COVID-19 treatment options in the face of a changing variant landscape.

**Competing Interest Statement:** Authors were employees of Invivyd, Inc. (A.H., K.N., I.Y., M.W.) or contractors of Invivyd, Inc. (L.H.) at the time this research was conducted and may hold stock or shares in Invivyd, Inc.

**Funding Statement:** This work was supported by Invivyd, Inc.

**Ethics and Reporting:** The authors declare that all relevant ethical guidelines have been followed, all necessary IRB and/or ethics committee approvals have been obtained, all necessary patient/participant consent has been obtained and the appropriate institutional forms archived for referenced trials supported by Invivyd, Inc.

## Introduction

Since the emergence of COVID-19 in 2020, the SARS-CoV-2 virus continues to demonstrate an ability to mutate and evade immunosuppression. Despite the widespread availability of vaccines and antiviral therapies, COVID-19 continues to be one of the leading causes of morbidity and mortality, particularly among immunocompromised individuals. In 2023, COVID-19 was the 10^th^ leading cause of death in the United States (CDC 2023), and persistent complications associated with post-acute sequalae continues to be recognized (CDC 2025b), demonstrating the critical need for an efficient accelerated path to introduce additional effective treatments.

In humans, antibody responses to SARS-CoV-2, especially high affinity IgG1 subtypes, emerge days after onset of symptoms and are associated with rapid reduction of viral load once present. High viral load is an independent predictor of acute COVID-19 severity and mortality, making treatment of infection via pharmaceutical monoclonal antibody (mAb) an attractive therapeutic strategy deeply rooted in native human immunobiology.

SARS-CoV-2 receptor-binding domain (RBD)-directed mAbs have established efficacy in the treatment of COVID-19, demonstrating 51-87% relative risk reduction in COVID-19–related hospitalization or all-cause death in clinical trials (Table 1) despite different target epitopes, pharmacokinetic profiles, and effector functionalities. While past mAbs proved an effective treatment, their availability was short lived as emergence of the Omicron variant and associated sublineages rendered them ineffective, resulting in deauthorization by the FDA. Antivirals are available as treatment options but have limitations such as drug-drug interactions (nirmatrelvir/ritonavir), renal dosing concerns (remdesivir), marginal clinical efficacy (molnupiravir), and universal lengthy duration of dosing, leaving a critical gap in patient care especially in high-risk and immunocompromised patients (Gilead Sciences 2024; Merck & Co 2024; Pfizer 2025).

**Table 1:**
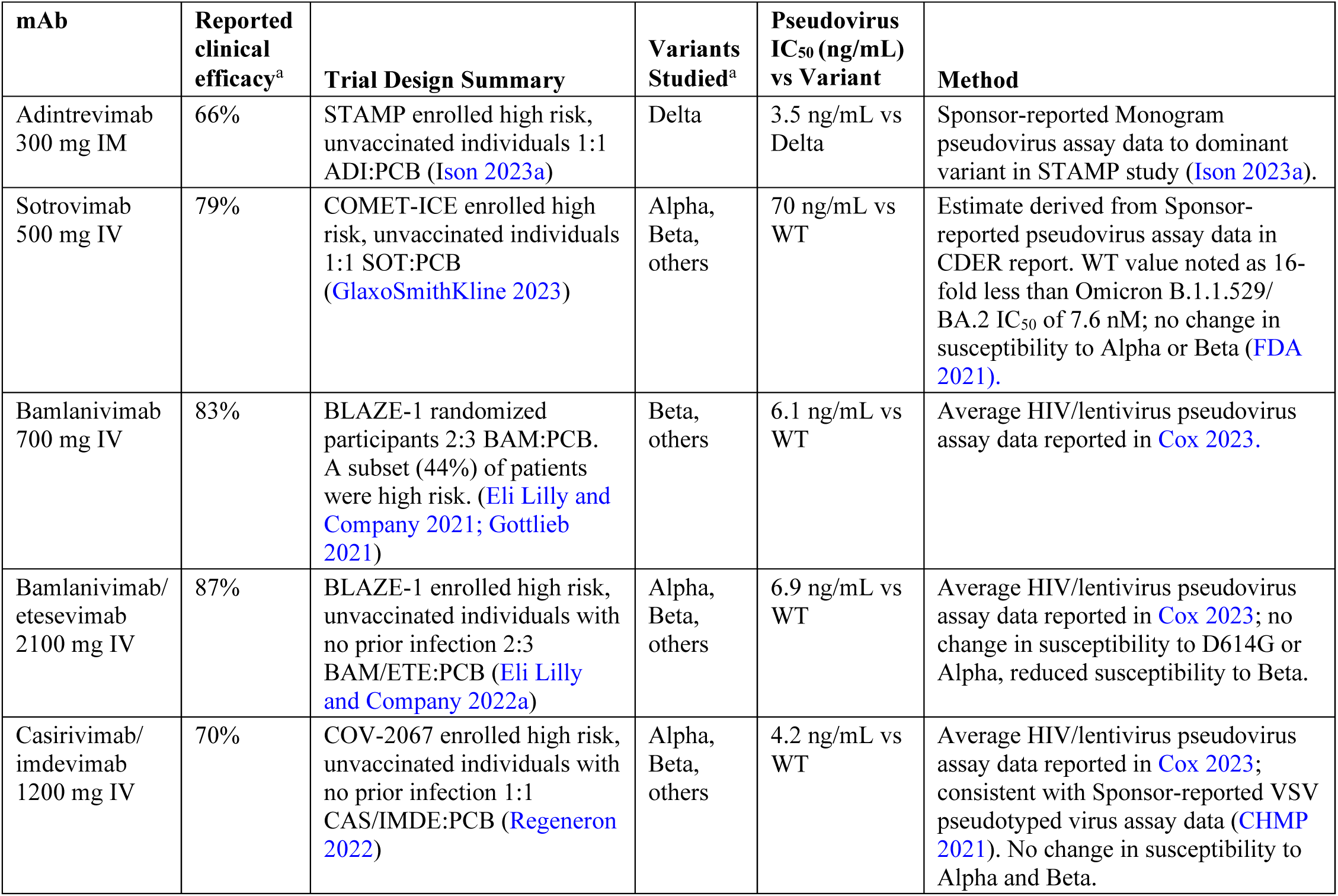
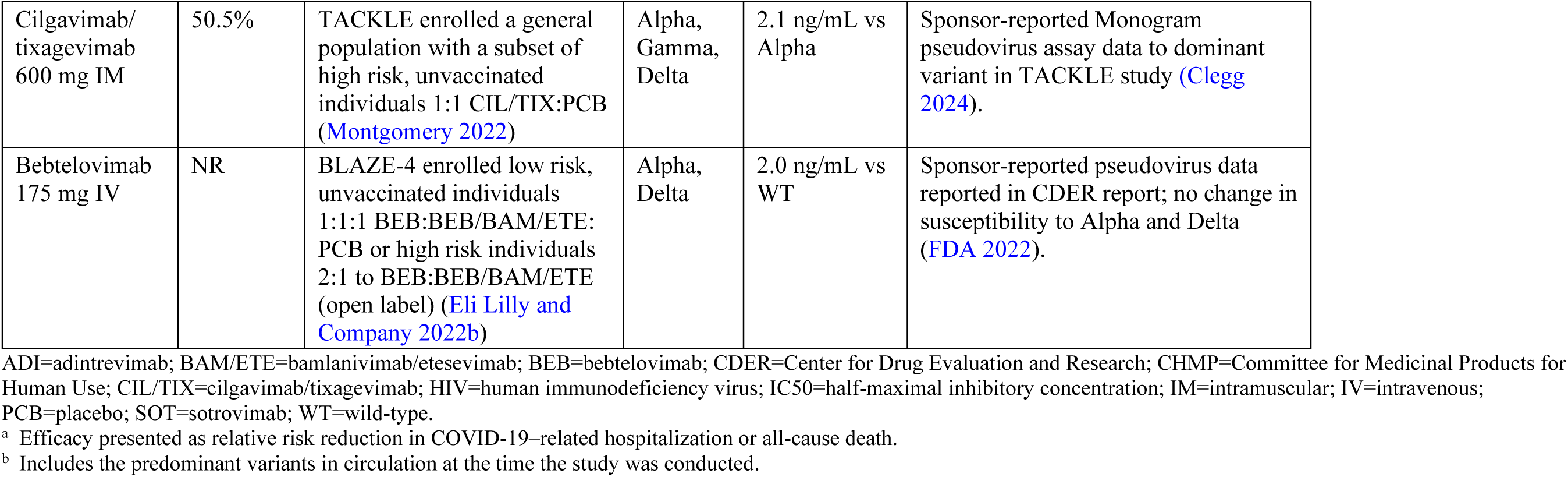
Summary of Half-maximal Inhibitory Concentration and Efficacy Against Circulating SARS-CoV-2 Variants of Historical mAbs.

| mAb | Reported clinical efficacy<sup>a</sup> | Trial Design Summary | Variants Studied<sup>a</sup> | Pseudovirus IC<sub>50</sub> (ng/mL) vs Variant | Method |
| --- | --- | --- | --- | --- | --- |
| Adintrevimab<br>300 mg IM | 66% | STAMP enrolled high risk, unvaccinated individuals 1:1 ADI:PCB (Ison 2023a) | Delta | 3.5 ng/mL vs Delta | Sponsor-reported Monogram pseudovirus assay data to dominant variant in STAMP study (Ison 2023a). |
| Sotrovimab<br>500 mg IV | 79% | COMET-ICE enrolled high risk, unvaccinated individuals 1:1 SOT:PCB (GlaxoSmithKline 2023) | Alpha, Beta, others | 70 ng/mL vs WT | Estimate derived from Sponsor-reported pseudovirus assay data in CDER report. WT value noted as 16-fold less than Omicron B.1.1.529/BA.2 IC<sub>50</sub> of 7.6 nM; no change in susceptibility to Alpha or Beta (FDA 2021). |
| Bamlanivimab<br>700 mg IV | 83% | BLAZE-1 randomized participants 2:3 BAM:PCB. A subset (44%) of patients were high risk. (Eli Lilly and Company 2021; Gottlieb 2021) | Beta, others | 6.1 ng/mL vs WT | Average HIV/lentivirus pseudovirus assay data reported in Cox 2023. |
| Bamlanivimab/<br>etesevimab<br>2100 mg IV | 87% | BLAZE-1 enrolled high risk, unvaccinated individuals with no prior infection 2:3 BAM/ETE:PCB (Eli Lilly and Company 2022a) | Alpha, Beta, others | 6.9 ng/mL vs WT | Average HIV/lentivirus pseudovirus assay data reported in Cox 2023; no change in susceptibility to D614G or Alpha, reduced susceptibility to Beta. |
| Casirivimab/<br>imdevimab<br>1200 mg IV | 70% | COV-2067 enrolled high risk, unvaccinated individuals with no prior infection 1:1 CAS/IMDE:PCB (Regeneron 2022) | Alpha, Beta, others | 4.2 ng/mL vs WT | Average HIV/lentivirus pseudovirus assay data reported in Cox 2023; consistent with Sponsor-reported VSV pseudotyped virus assay data (CHMP 2021). No change in susceptibility to Alpha and Beta. |
| Cilgavimab/<br>tixagevimab<br>600 mg IM | 50.5% | TACKLE enrolled a general population with a subset of high risk, unvaccinated individuals 1:1 CIL/TIX:PCB (Montgomery 2022) | Alpha, Gamma, Delta | 2.1 ng/mL vs Alpha | Sponsor-reported Monogram pseudovirus assay data to dominant variant in TACKLE study (Clegg 2024). |
| Bebtelovimab<br>175 mg IV | NR | BLAZE-4 enrolled low risk, unvaccinated individuals 1:1:1 BEB:BEB/BAM/ETE:PCB or high risk individuals 2:1 to BEB:BEB/BAM/ETE (open label) (Eli Lilly and Company 2022b) | Alpha, Delta | 2.0 ng/mL vs WT | Sponsor-reported pseudovirus data reported in CDER report; no change in susceptibility to Alpha and Delta (FDA 2022). |

ADI=adintrevimab; BAM/ETE=bamlanivimab/etesevimab; BEB=bebtelovimab; CDER=Center for Drug Evaluation and Research; CHMP=Committee for Medicinal Products for Human Use; CIL/TIX=cilgavimab/tixagevimab; HIV=human immunodeficiency virus; IC50=half-maximal inhibitory concentration; IM=intramuscular; IV=intravenous; PCB=placebo; SOT=sotrovimab; WT=wild-type.
<sup>a</sup> Efficacy presented as relative risk reduction in COVID-19–related hospitalization or all-cause death.
<sup>b</sup> Includes the predominant variants in circulation at the time the study was conducted.

Rapid virus evolution challenges traditional development pathways, as non-susceptible variants can emerge faster than development and regulatory review of mAbs can occur. Fortunately, candidate mAb antiviral activity across viral variants can be measured and compared via clinical serum virus neutralizing antibody titers that correlate to demonstrated clinical outcomes from predicate mAbs, creating a surrogate marker and analytic immunobridging approach useful for rapid assessment of novel mAb efficacy, especially for mAbs engineered from a clinically established molecular ancestor.

Immunobridging is a powerful tool for advancing timely development of treatments for infectious diseases and has been widely used in the development of vaccines to enable rapid vaccine composition evolution and regulatory review. In the context of emerging viral threats with fluctuating transmission dynamics like COVID-19, immunobridging offers a scientifically grounded and efficient alternative to large-scale efficacy trials that could be routinely deployed to accelerate mAb development, while also describing the pharmaceutical amplification of the canonical, native human antibody response to acute SARS-CoV-2 infection. It enables assessment of treatment performance across different populations (eg, children, immunocompromised individuals) without awaiting natural disease exposure. When supported by strong mechanistic evidence and clinical validation, immunobridging can provide a strong foundation for accelerated development, regulatory decision making, and ultimately, more timely patient access to potentially life-saving interventions.

Pemivibart, a potent SARS-CoV-2 RBD-directed mAb, offers a rational solution for patients with unmet need. Pemivibart was engineered from its parent molecule adintrevimab, which previously demonstrated efficacy in both treatment of high-risk ambulatory patients with COVID-19 (STAMP trial) and pre- and post-exposure prophylaxis of COVID-19 (EVADE trial) against the Delta variant (Ison 2023a; Ison 2023b). In the STAMP trial (NCT04805671), a single 300 mg dose of adintrevimab was administered intramuscularly (IM) to participants with at least one risk factor for severe disease who were diagnosed with COVID-19. The proportion of participants with COVID-19–related hospitalization or all-cause death (primary efficacy endpoint) was 4.7% (8/169) in the adintrevimab arm compared with 13.8% (23/167) in the placebo arm, a 66% relative risk reduction (P=0.0047)(Ison 2023a). Adintrevimab lost activity upon emergence of the Omicron BA.2 variant and development was suspended.

Pemivibart demonstrates broad and robust neutralizing activity against historical and emerging SARS-CoV-2 variants, including all Omicron sublineages evaluated by Invivyd to date (Invivyd 2025). On March 22, 2024, the FDA granted Emergency Use Authorization (EUA) for pemivibart for pre-exposure prophylaxis of COVID-19 in moderately to severely immunocompromised adults and adolescents, based on immunobridging data from the phase 3 CANOPY trial (Schmidt 2024; Invivyd 2025; Wolfe 2025). In this trial (NCT06039449), immunocompromised participants received 4500 mg of pemivibart intravenously on Day 1 and Month 3, with surrogate efficacy based on neutralizing antibody titers (Schmidt 2024). Clinical efficacy of pemivibart was later demonstrated in the CANOPY trial in a non-immunocompromised placebo-controlled cohort with relative risk reduction in symptomatic COVID-19 of 84% through 6 months, validating immunobridging as a reliable predictor of mAb effectiveness (Wolfe 2025).

We therefore applied a similar immunobridging framework to evaluate pemivibart in the treatment of acute COVID-19 in an effort to accelerate treatment availability for patients with an urgent unmet need who remain vulnerable despite vaccination and antiviral therapies.

Immunobridging analyses revealed that the predicted efficacy of pemivibart is likely equivalent to that of prior COVID-19 mAb therapeutics with demonstrated clinical efficacy and that antiviral neutralization is comparable to adintrevimab, the prototype antibody from which pemivibart was developed (Ison 2023a). However, we failed to secure FDA authorization of pemivibart for treatment of COVID-19. Nonetheless, a surrogate development approach and the U.S. regulatory response to our initial approach provides a road map for advancing antiviral treatment via mAb therapy for SARS-CoV-2 globally, and possibly for additional viral or microbial diseases in the future.

## Methods

For treatment of COVID-19, an immunobridging approach for pemivibart against contemporary variants, along with safety data from the previously described CANOPY clinical trial, was applied (Schmidt 2024; Wolfe 2025). The immunobridging analysis used three complementary methods: Method 1 employed a comparison of calculated neutralizing antibody titers for pemivibart versus adintrevimab based on population pharmacokinetics (popPK) modeling; Method 2 compared calculated neutralizing antibody titers of pemivibart to other SARS-CoV-2– directed mAbs historically used for treatment (but no longer effective) based on popPK modeling; and Method 3 compared the clinical efficacy and neutralizing dose of comparator mAbs versus pemivibart based on a published meta-analysis (Stadler 2023).

Selected relevant variants evaluated for pemivibart immunobridging include the JN.1-lineage variants JN.1, KP.3.1.1, XEC, and LP.8.1. These four variants encompass the range of available IC_50_ values against the most prevalent variants in circulation since 2024 (CDC 2025a) (Table 2). Although no longer in circulation, JN.1 was the primary variant circulating during the CANOPY trial at the time of the pemivibart EUA application for COVID-19 prevention; KP.3.1.1 and XEC were dominant variants circulating during the CANOPY trial; and LP.8.1 is the most current dominant variant as per the CDC variant tracker for which data is available. Evaluation of the LP.8.1 variant was not included in the EUA application for treatment (not available at the time of submission).

**Table 2:**
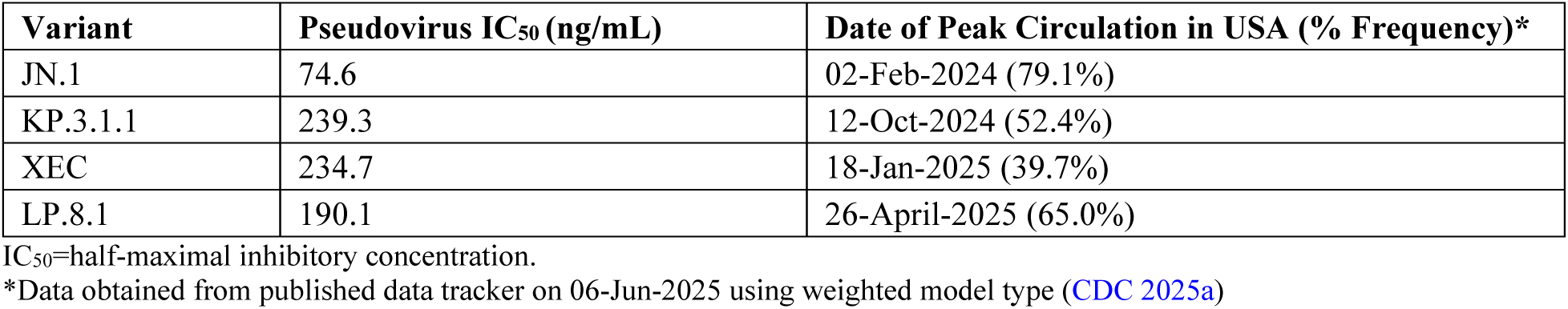
Relevant Dominant Variants Evaluated for Immunobridging of Pemivibart for Treatment Indication.

| Variant | Pseudovirus IC<sub>50</sub> (ng/mL) | Date of Peak Circulation in USA (% Frequency)* |
| --- | --- | --- |
| JN.1 | 74.6 | 02-Feb-2024 (79.1%) |
| KP.3.1.1 | 239.3 | 12-Oct-2024 (52.4%) |
| XEC | 234.7 | 18-Jan-2025 (39.7%) |
| LP.8.1 | 190.1 | 26-April-2025 (65.0%) |

IC<sub>50</sub>=half-maximal inhibitory concentration.
\*Data obtained from published data tracker on 06-Jun-2025 using weighted model type (CDC 2025a)

In Immunobridging Method 1, an immunobridging analysis comparing the neutralizing antibody titers (GMTs) of pemivibart to adintrevimab was employed. Adintrevimab was particularly potent against the Delta variant, producing titers that were several-fold higher than those expected to produce maximal efficacy. This was recognized as a possible limitation for immunobridging to adintrevimab, as the pemivibart titers required for successful immunobridging were also likely inflated above those required for maximal efficacy.

For this analysis, GMTs, calculated as estimated average drug concentrations from dosing to specified times of interest (area under the curve [AUC]/time obtained with popPK analysis) divided by pseudovirus neutralization assay half-maximal inhibitory concentration (IC_50_) against a relevant variant, were used to account for the difference in the route of administration for pemivibart (intravenous [IV]) compared with adintrevimab (IM). Adintrevimab titer data were obtained from a Phase 1 trial and the STAMP trial during the Delta variant period and compared to pemivibart titers against selected current variants at early timepoints following onset of symptoms. Strict immunobridging is established if the lower bound of the 90% confidence interval (CI) of the estimated geometric mean ratio (GMR) of GMTs from pemivibart compared with adintrevimab is 0.8 or higher.

In the STAMP trial, the most significant treatment effects were observed if dosing occurred in the first 3 days after symptom onset, and the largest impact to viral load was achieved by day 5, suggesting immunobridging through 5 days is a reasonable target for predicting efficacy (Ison 2023a). Although mechanisms of actions may differ, sustained antiviral activity through a maximum of 5 days is also the intended target for several small molecule products for treatment of COVID-19 (ie, nirmatrelvir/ritonavir, remdesivir, molnupiravir) (Gilead Sciences 2024; Merck & Co 2024; Pfizer 2025).

In Immunobridging Method 2, GMTs based on estimated average concentration (AUC/time) of pemivibart against relevant variants following a single IV dose were compared with GMTs of other mAbs previously shown to be efficacious for treatment of COVID-19 against variants circulating at the time trials were conducted. Briefly, a pemivibart popPK model and publicly available popPK models of comparator anti-COVID-19 mAbs were used to simulate individual antibody concentration versus time profiles in over 2400 virtual subjects using demographic information from an adintrevimab Phase 3 study (Chigutsa 2021; Dougan 2022; Sager 2023; Clegg 2024; Lin 2024). Geometric mean of average concentration profile of each antibody was then calculated and derived to obtain GMTs, where IC_50_ values for comparator mAbs were derived from sponsor-reported data or alternative sources using pseudovirus neutralization assay (Table 1) (CHMP 2021; FDA 2021; FDA 2022; Cox 2023; Clegg 2024).

Immunobridging Method 3 compared the clinical efficacy and neutralizing dose of comparator mAbs versus pemivibart. The dose-response relationship between neutralizing dose and efficacy developed by Stadler et al (Stadler 2023) provides a direct quantitative means of assessing potential therapeutic efficacy based on in vitro neutralization data and mAb dose.

## Results

### Immunobridging Method 1

Immunobridging from pemivibart to adintrevimab against the Delta variant was nominally established for selected circulating variants: through 4 days for variants KP.3.1.1 and XEC, through 6 days for LP.8.1, and through >14 days for JN.1 (Figure 1 and Table 3). For KP.3.1.1 and XEC, pemivibart GMTs remained at 77% and 79% of adintrevimab GMTs at the immunobridging target of 5 days, respectively. Average titers against all selected variants remained high and stable through 14 days (3662-11,748), suggesting pemivibart likely continues to provide long-acting antiviral benefit well beyond the acute infection phase, although absolute titers fluctuate depending on variant analyzed.

**Figure 1:**
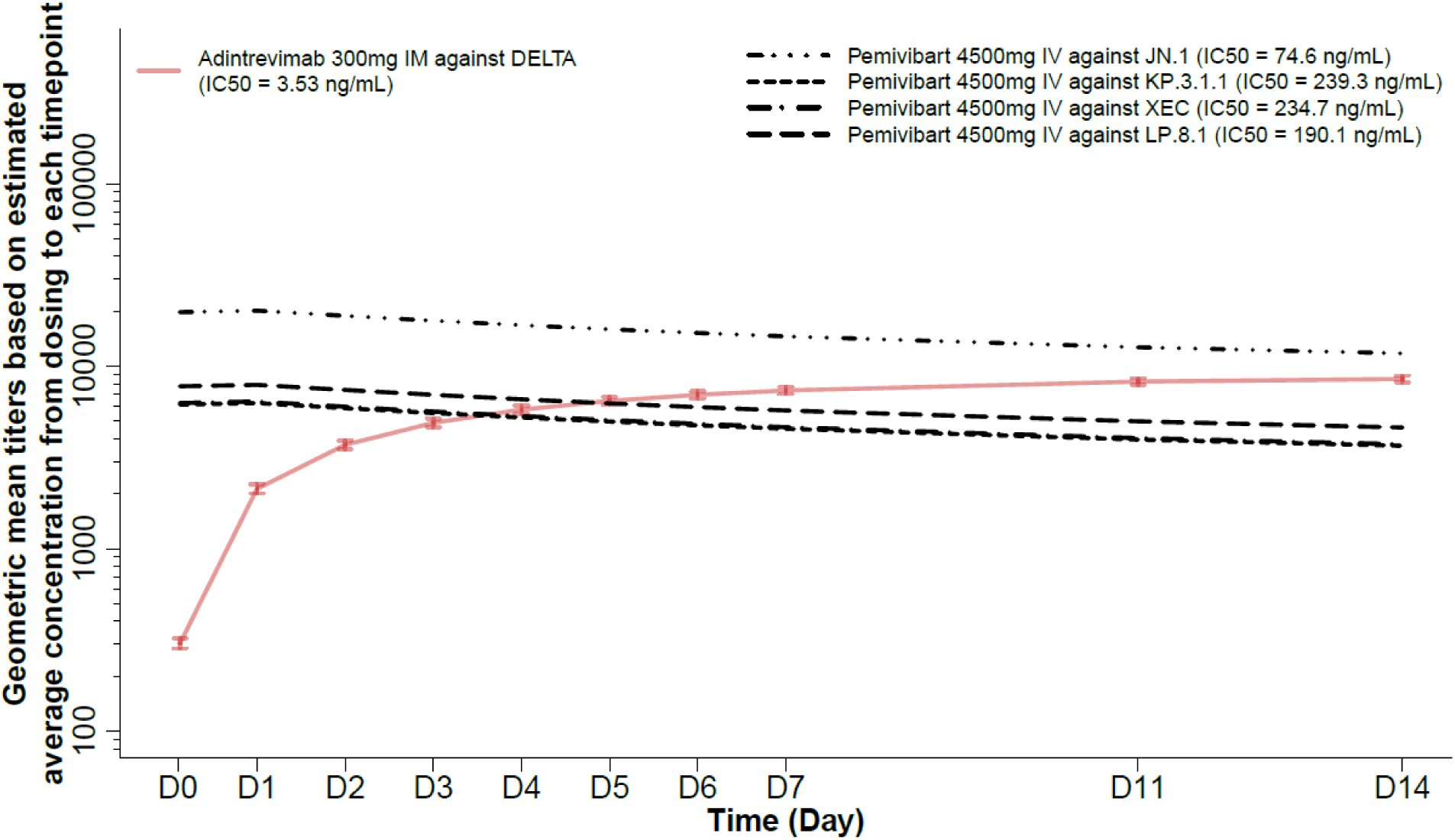
Pemivibart Treatment Immunobridging to Adintrevimab (Method 1) CI=confidence interval; GMT=geometric mean titer; IC_50_=half-maximal inhibitory concentration. GMT and 90% CI at each time point for pemivibart and adintrevimab were summarized from population PK model−estimated post hoc concentrations of individual subjects in correspondent pemivibart phase 3 study (VYD222−PREV−001) and adintrevimab phase 3 study (ADG20−TRMT−001), respectively. Concentrations were then divided by correspondent IC_50_ values of dominant circulating variants when trials were conducted as described in Table 2 for pemivibart and Table 1 for adintrevimab.

**Table 3:**
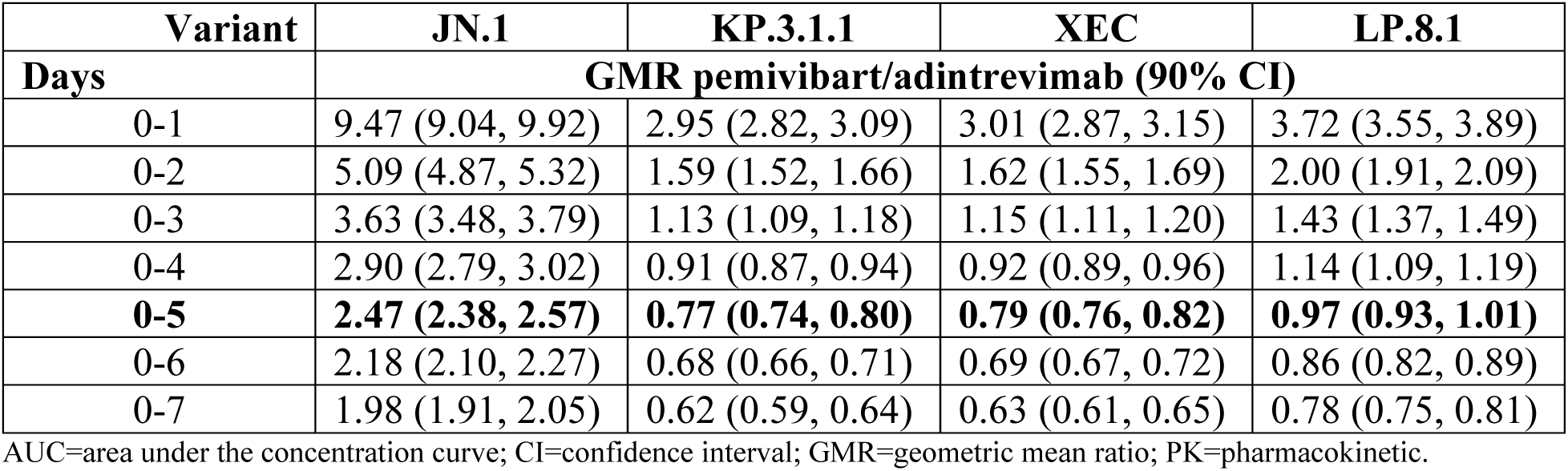
Summary of Geometric Mean Ratios of Average Concentration (AUC/time) of Pemivibart Against Select SARS-CoV-2 Variants to Adintrevimab Against Delta by Pseudotyped Virus Assay Based on Population PK Estimate.

### Immunobridging Method 2

Pemivibart GMTs to selected variants are between those of sotrovimab and bebtelovimab, with neutralizing titers approximately 4-12 fold higher than those provided by a single 500 mg IV dose of sotrovimab through 14 days post dose (Figure 2). Sotrovimab provided clinically meaningful efficacy for the treatment of COVID-19 with a 79% reduction in COVID-19–related hospitalization or all-cause death, respectively (Table 1)(GlaxoSmithKline 2023).

**Figure 2:**
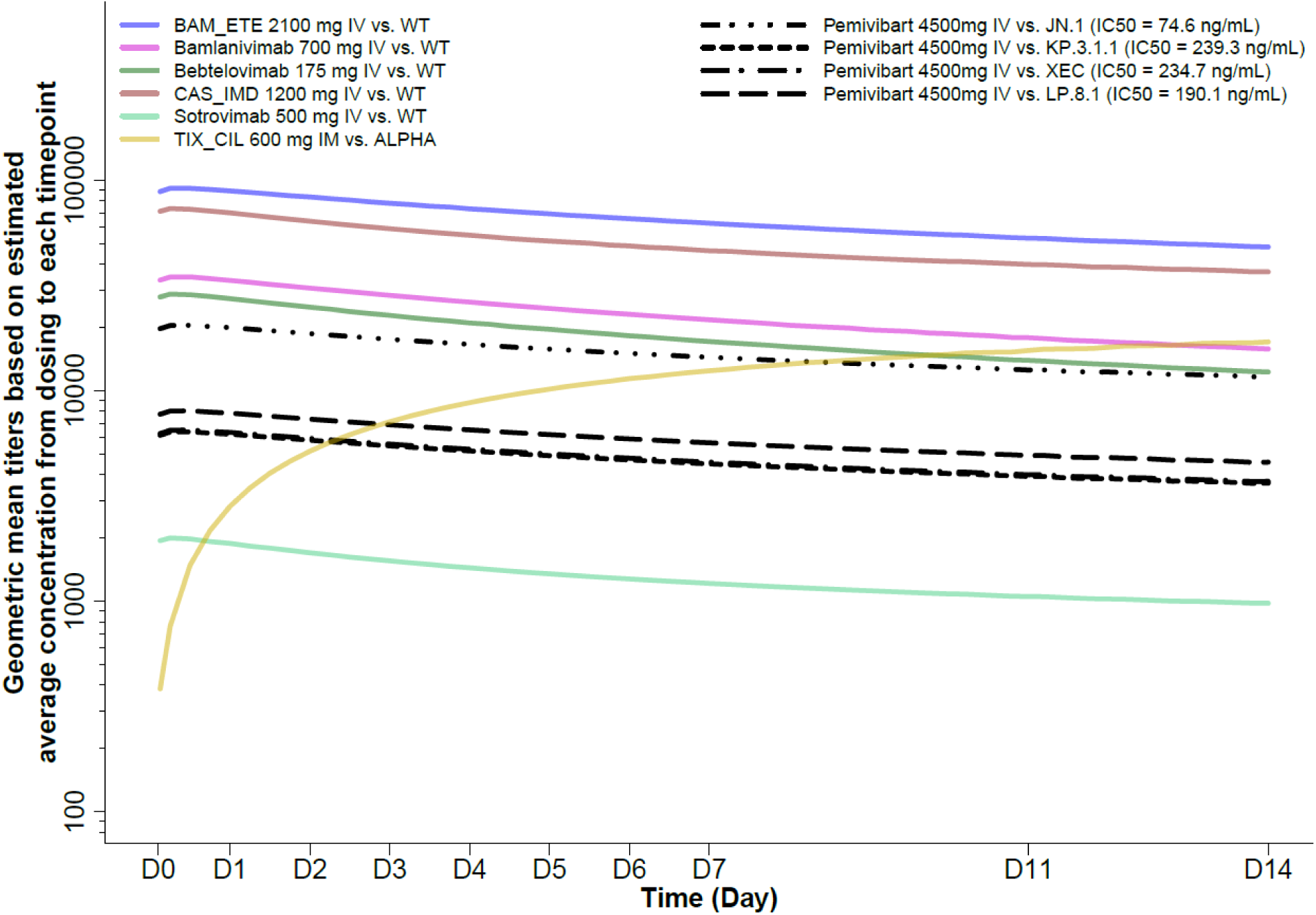
Pemivibart Treatment Immunobridging to Comparator mAbs (Method 2) BAM_ETE=bamlanivimab and etesevimab; CAS_IMD=casirivimab and imdevimab; IC_50_=half-maximal inhibitory concentration; IM=intramuscular; IV=intravenous; PK=pharmacokinetic; TIX_CIL=tixagevimab and cilgavimab; VYD222=pemivibart; WT=wild-type virus. The PK profiles for comparator mAbs were simulated using population PK parameters available in the public domain. PK profiles for pemivibart are based on the pemivibart popPK model. Titers were calculated based on published IC_50_ data summarized in Table 1.

### Immunobridging Method 3

The range of neutralizing doses observed across the specific doses of mAbs with prior EUA for the treatment of COVID-19 is between 4.4 (sotrovimab 500 mg) and 123.8 (bamlanivimab/etesevimab 2.1 g) (Figure 3). Upon evaluation of pemivibart 4500 mg IV, the neutralizing titers against selected SARS-CoV-2 variants were 11.2 to 35.8, which falls on the plateau of the efficacy curve. The neutralizing dose of pemivibart is at least 2.5 times greater than sotrovimab for the least susceptible variants KP.3.1.1 and XEC and above the range observed across trials of convalescent plasma (0.1 to 1.5), which is authorized for emergency use for treatment of COVID-19 in individuals with immune compromise.

**Figure 3:**
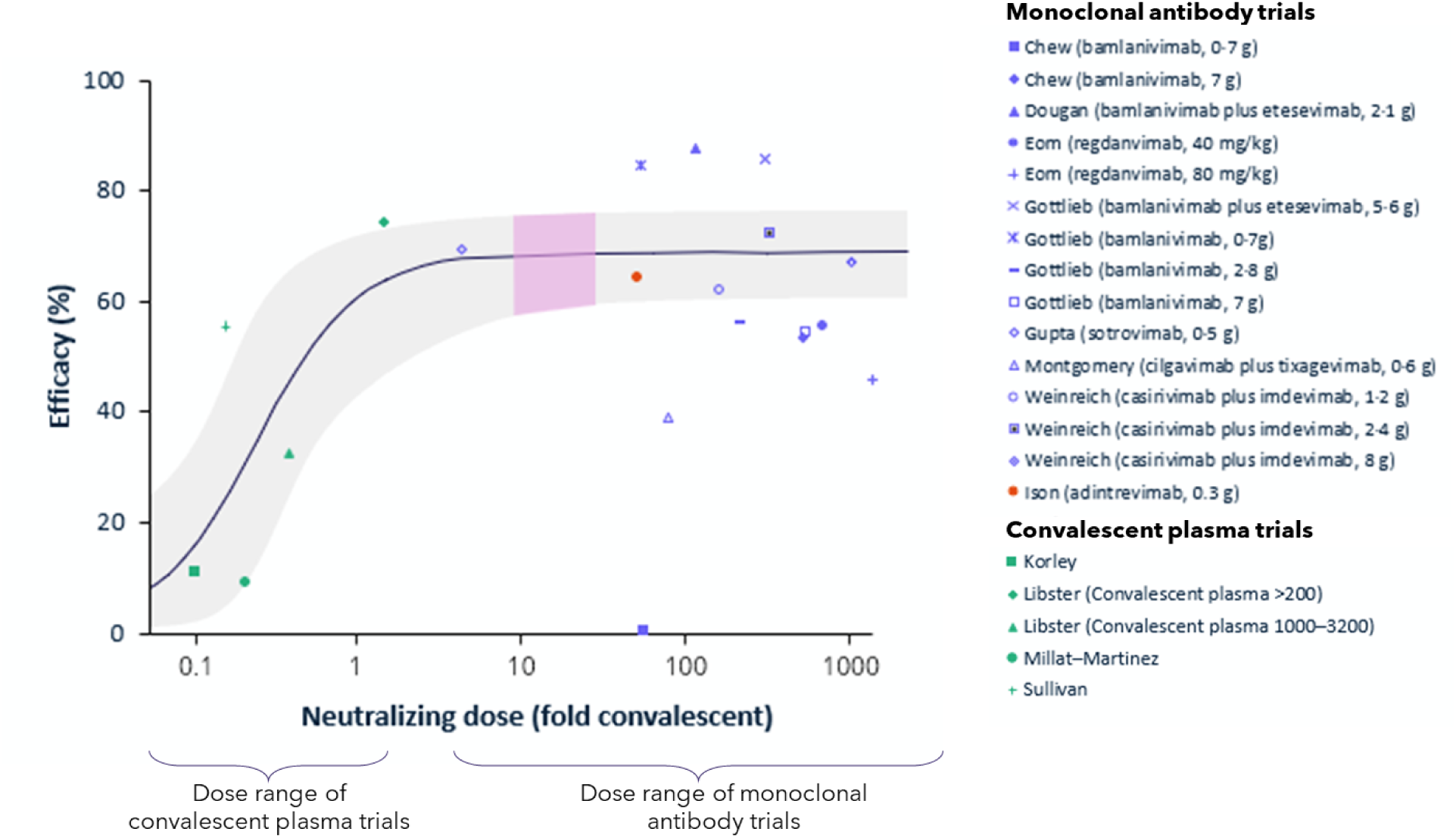
Pemivibart Treatment Immunobridging Meta-Analysis (Method 3) A logistic model was fitted to the data on efficacy and corresponding neutralising dose for each study as described in (Stadler 2023). The fitted curve allows for maximum efficacy (estimated 67.5%). The gray shaded area indicates the 95% CI. The pink shaded area represents the range of neutralizing titers (∼11.2 to 35.8) against SARS-CoV-2 variants JN.1, KP.3.1.1, XEC, and LP.8.1 following a single dose of pemivibart 4500 mg IV. The yellow bullet represents adintrevimab neutralizing dose (50.9) based on IC_50_ against pseudovirus. Figure adapted from (Stadler 2023).

It is important to note that based on the meta-analysis from Stadler et al (Stadler 2023), the efficacy rate of all mAbs plateau once a neutralizing dose is reached. Adintrevimab, along with other mAbs authorized for treatment, falls on the plateau of the efficacy curve. These mAbs have significantly higher neutralizing doses than sotrovimab, but without increased efficacy. By comparison, 500 mg sotrovimab IV is at the threshold of plateaued maximal efficacy among the comparators against correspondent dominant virus variants. Hence, clinical equivalence of neutralizing titers of pemivibart to other mAbs is established in this model, where an effective neutralizing dose is present on the plateau of the dose-response curve.

## Discussion

Immunobridging has been a commonly accepted pathway to facilitate approval of vaccines. Pemivibart was the first SARS-CoV-2 mAb to effectively utilize the immunobridging approach to obtain EUA for prevention of COVID-19 prior to the availability of clinical efficacy data (Schmidt 2024). In the prevention immunobridging analysis, strict immunobridging (lower bound of 90% CI >0.8) of pemivibart to adintrevimab through 3 months in an immunocompromised population was not met against the JN.1 variant dominant at the time of the analysis. However, the neutralizing titer levels of pemivibart were in line with other previous mAbs with established efficacy in the prevention setting, leading to authorization based on totality of evidence and demonstrating the value of using multiple methods for immunobridging (Schmidt 2024).

Since then, pemivibart has demonstrated robust clinical efficacy in the prevention of RT-PCR-confirmed symptomatic COVID-19 in an immunocompetent placebo-controlled population through 3 and 6 months, with a standardized relative risk reduction (sRRR) of 94% (p=0.009) and 84% (p<0.001), respectively, following 2 doses of pemivibart at a 3-month interval in the CANOPY trial. Pemivibart also continued to show clinical efficacy (sRRR of 74%; p<0.001) against relevant circulating variants through 12 months, despite having waning titer levels, suggesting that while immunobridging is an extremely useful tool for predicting protection, immunobridging target titers do not necessarily define the lower bound at which clinical efficacy is actually present (Wolfe 2025).

A similar immunobridging approach was taken to support the use of pemivibart in the treatment of COVID-19. Using neutralizing data against recent dominant variants, pemivibart demonstrated immunobridging through three different methods, all of which demonstrate antiviral activity in acute COVID-19 that would accelerate and amplify the native antibody response. Strict immunobridging to adintrevimab was achieved through the first 4 days of the treatment period for XEC and KP.3.1.1, through 6 days for LP.8.1, and through >14 days for JN.1 (Figure 1). A time period of 5 days from symptom onset has been demonstrated to be most critical in COVID-19 treatment, given clear observations that early intervention and virologic reductions within the first 5 days are highly correlated with clinical efficacy (Abusalem 2021; Montgomery 2022; Elias 2024). Data from the adintrevimab treatment trial (STAMP) showed that an intramuscular injection of adintrevimab, despite failing to generate maximal titer levels until Day 7-10, correlated to a 66% reduction in hospitalization or death with the greatest impact on viral load reduction observed at day 5 post-treatment (Ison 2023a). The pharmacokinetic profile of pemivibart administered intravenously lends itself to providing immediate and superior titer levels compared to adintrevimab for the first 3 days after dosing and comparable titer levels though at least 5 days (Figure 1, Table 3), supporting meaningful clinical potential. Although mechanisms of actions may differ, sustained antiviral activity through a maximum of 5 days is also the intended target for several small molecule products for treatment of COVID-19. Achieving maximal titers immediately post-IV treatment has likely advantages of abolishing clinically significant viremia more quickly and warding against symptom breakthrough or untoward COVID-19 sequalae than other routes of administration.

In comparing pemivibart to other mAbs with known clinical efficacy, neutralizing titer levels of pemivibart most notably remained above the titers of sotrovimab for the full analysis period for all variants tested (>14 days; Figure 2). Likewise, the neutralizing dose of pemivibart falls on the plateau of the dose-response curve generated by Stadler et al (Stadler 2023), establishing clinical equivalence of pemivibart to other mAbs in this model (Figure 3). The totality of the data immunobridging pemivibart to adintrevimab and other comparator mAbs for COVID-19 treatment is similar to the package leading to EUA for pemivibart in COVID-19 prevention, with an additional approximately 9 months of safety data from CANOPY and post-marketing safety data available.

The results of the analysis were submitted to the FDA for EUA of pemivibart for the treatment of mild to moderate COVID-19. The agency indicated uncertainties in the analysis and were therefore unable to conclude a favorable benefit-risk profile, despite the consistent results from the multi-pronged immunobridging methodologies. Specifically, minimum titers required for successful treatment and the duration of time that titers must be present in the immune compromised have not been defined due to a scarcity of placebo-controlled trials in this high-risk population. In the CANOPY study, neutralizing titers calculated from serum pemivibart concentrations in immunocompromised participants were similar to immunocompetent participants; however, all prior mAb EUAs for treatment were based on studies in immunocompetent participants. Clinical efficacy with pemivibart in prevention has also demonstrated that antiviral activity is present at much lower titers than were evaluated in any studies with historical mAbs and may also apply to treatment as viral neutralization is a key mechanism of action for both indications. The duration of neutralization required in the immune compromised may also be longer due to lack of viral clearance by a faulty or suppressed immune system, although studies have clearly demonstrated the critical role of early intervention and early viral load reduction in preventing disease progression. Placebo-controlled studies in the immune compromised that demonstrate true titer thresholds of clinical effectiveness and duration for treatment are the most straightforward way to definitively answer this question.

Another uncertainty raised by the FDA suggested immunobridging to comparator mAbs with variability in Fc effector function may contribute to efficacy beyond viral neutralization and therefore may represent a possible inconsistency in our approach. Pemivibart has intact Fc effector functions, similar to most other historical mAbs for treatment of COVID-19, with the exception of tixagevimab/cilgavimab and etesevimab (Edgar 2024). Although overall efficacy conclusions may not be overtly impacted, the most appropriate direct immunobridging comparison may to be mAbs developed using the same platform and structural backbone (eg, pemivibart to adintrevimab with only 8 amino acid differences). Importantly, however, the dose-response immunobridging analysis suggests all mAbs with neutralizing dose equal or greater to sotrovimab should have generally equivalent efficacy, as the maximal efficacy threshold has been reached – which may diminish the importance of showing neutralizing titers reach a specific numeric level independently or in comparison to other historical mAbs. Here, the totality of evidence seems to favor a reasonable expectation of effect of pemivibart in treatment of COVID-19. The safety profile of pemivibart, supported by the CANOPY Phase 3 trial, indicates manageable risks, with infusion-related reactions (6%) and rare anaphylaxis (0.6%) across immune compromised and immune competent populations mitigated by standard monitoring (Invivyd 2025). Therefore, the risk-benefit of pemivibart for the most at-risk populations with unmet need would support its use in COVID-19 treatment.

Immunocompromised patients face unique challenges upon SARS-CoV-2 exposure. Current vaccines are insufficient to protect immunocompromised individuals from infection, resulting in high vulnerability to severe outcomes (Britton 2022; Ferdinands 2022; Evans 2023; Ku 2023; Link-Gelles 2025). Indeed, studies demonstrate COVID-19 hospitalization, intensive care unit admission, and mortality rates were approximately 6-11 fold higher in immunocompromised populations (Evans 2023). The inability to mount a proper immune response may also result in prolonged viral shedding, prolonging the acute complications of COVID-19 (Kang 2023).

Access to standard antiviral therapies is limited due to drug-drug interactions with nirmatrelvir/ritonavir and the logistical barriers of multi-day remdesivir infusions for some individuals (IDSA 2025). Immunobridging analyses supporting pemivibart utility in treatment, as a single-dose IV therapy with no known drug-drug interactions that mechanistically amplifies and precedes the native human antibody response to acute infection, could address a critical gap in care for populations who remain at high risk of severe COVID-19 outcomes.

## Conclusion

We demonstrate a positive risk-benefit profile of pemivibart by using multiple complementary immunobridging methods for treatment of COVID-19. Residual clinical uncertainty embedded into any immunobridging exercise is minimized via the consistency and clear clinical utility of sVNA titer analysis. Establishment of defined authorization or approval pathways based on such immunobridging for rapid development and approval of COVID-19 mAbs could allow sick populations and their care teams to access substantial immunologic support in the form of mAb therapy that further lowers the burden of COVID-19 disease in America. This development approach, which draws on quantitative virology and natural biological mechanisms, may be suitable to support authorization of future mAbs for COVID-19 and may have broader application in viral diseases where a rapidly evolving variant landscape necessitates accelerated novel treatments for those at risk of severe disease.

## Data Availability

all data produced in the present study are available upon reasonable request to the authors

## References

1. Abusalem L, Wood C, Crescencio JCR, et al. (2021). 519. Risk Factor Analysis for Hospital Admission Following Severe Acute Respiratory Syndrome Coronavirus-2 (SARS-CoV-2) Monoclonal Antibody Treatment. Open Forum Infect Dis. 8:S361.

2. Britton A, Embi PJ, Levy ME, et al. (2022). Effectiveness of COVID-19 mRNA vaccines against COVID-19-associated hospitalizations among immunocompromised adults during SARS-CoV-2 Omicron predominance-VISION Network, 10 States, December 2021-August 2022. MMWR Morb Mortal Wkly Rep. 71(42):1335–1342. doi:10.15585/mmwr.mm7142a4

3. CDC. (2023). Leading Causes of Death. https://www.cdc.gov/nchs/fastats/leading-causes-of-death.htm CDC. (2025a). COVID Data Tracker. Variant proportions. https://covid.cdc.gov/covid-data-tracker/#variant-proportions

4. CDC. (2025b). Long COVID Basics. https://www.cdc.gov/covid/long-term-effects/index.html

5. Chigutsa E, O’Brien L, Ferguson-Sells L, et al. (2021). Population Pharmacokinetics and Pharmacodynamics of the Neutralizing Antibodies Bamlanivimab and Etesevimab in Patients With Mild to Moderate COVID-19 Infection. Clin Pharmacol Ther. 110(5):1302–1310. doi:10.1002/cpt.2420

6. CHMP. (2021). Ronapreve EPAR Public assessment report. Nov 19, 2021. https://www.ema.europa.eu/en/documents/assessment-report/ronapreve-epar-public-assessment-report_en.pdf

6. Clegg LE, Stepanov O, Schmidt H, et al. (2024). Accelerating therapeutics development during a pandemic: population pharmacokinetics of the long-acting antibody combination AZD7442 (tixagevimab/cilgavimab) in the prophylaxis and treatment of COVID-19. Antimicrob Agents Chemother. 68(5):e0158723. doi:10.1128/aac.01587-23

7. Cox M, Peacock TP, Harvey WT, et al. (2023). SARS-CoV-2 variant evasion of monoclonal antibodies based on in vitro studies. Nat Rev Microbiol. 21(2):112–124. doi:10.1038/s41579-022-00809-7

8. Dougan M, Azizad M, Chen P, et al. (2022). Bebtelovimab, alone or together with bamlanivimab and etesevimab, as a broadly neutralizing monoclonal antibody treatment for mild to moderate, ambulatory COVID-19. medRxiv. doi:10.1101/2022.03.10.22272100

9. Edgar JE, Bournazos S. (2024). Fc-FcgammaR interactions during infections: From neutralizing antibodies to antibody-dependent enhancement. Immunol Rev. 328(1):221–242. doi:10.1111/imr.13393

10. Eli Lilly and Company. (2021). Fact Sheet for Health Care Providers: Emergency Use Authorization of Bamlanivimab. Updated March 2021. https://www.fda.gov/media/143603/download

11. Eli Lilly and Company. (2022a). Fact Sheet for Health Care Providers: Emergency Use Authorization (EUA) of Bamlanivimab and Etesevimab. Updated January 24, 2022. https://www.fda.gov/media/145802/download

12. Eli Lilly and Company. (2022b). Fact Sheet for Health Care Providers: Emergency Use Authorization of Bebtelovimab. Updated November 2022. https://www.fda.gov/media/156152/download

13. Elias KM, Khan SR, Stadler E, et al. (2024). Viral clearance as a surrogate of clinical efficacy for COVID-19 therapies in outpatients: a systematic review and meta-analysis. Lancet Microbe. 5(5):e459–e467. doi:10.1016/S2666-5247(23)00398-1

14. Evans RA, Dube S, Lu Y, et al. (2023). Impact of COVID-19 on immunocompromised populations during the Omicron era: insights from the observational population-based INFORM study. The Lancet Regional Health – Europe. 35(100747). doi:10.1016/j.lanepe.2023.100747

15. FDA. (2021). Emergency Use Authorization (EUA) for Sotrovimab Center for Drug Evaluation and Research (CDER) Review. May 26, 2021. https://www.fda.gov/media/150130/download?attachment

16. FDA. (2022). Emergency Use Authorization (EUA) for Bebtelovimab (LY-CoV1404). Center for Drug Evaluation and Research (CDER) Review, Reference ID: 4936741. February 18, 2022. https://www.fda.gov/media/156396/download

17. Ferdinands JM, Rao S, Dixon BE, et al. (2022). Waning of vaccine effectiveness against moderate and severe covid-19 among adults in the US from the VISION network: test negative, case-control study. Bmj. 379:e072141. doi:10.1136/bmj-2022-072141

18. Gilead Sciences. (2024). Veklury (Remdesivir) Prescribing Information. Revised July 2024. https://www.gilead.com/-/media/files/pdfs/medicines/COVID-19/veklury/veklury_pi.pdf

19. GlaxoSmithKline. (2023). Fact Sheet for Healthcare Providers: Emergency Use Authorization for Sotrovimab. Updated March 2023. https://www.fda.gov/media/149534/download

20. Gottlieb RL, Nirula A, Chen P, et al. (2021). Effect of Bamlanivimab as Monotherapy or in Combination With Etesevimab on Viral Load in Patients With Mild to Moderate COVID-19: A Randomized Clinical Trial. Jama. 325(7):632–644. doi:10.1001/jama.2021.0202

21. IDSA. (2025). IDSA Guidelines on the Treatment and Management of Patients with COVID-19. https://www.idsociety.org/practice-guideline/covid-19-guideline-treatment-and-management/

22. Invivyd. (2025). Fact Sheet for Healthcare Providers: Emergency Use Authorization of PEMGARDA (Pemivibart). Updated May 2025. https://www.fda.gov/media/177067/download?attachment

23. Ison MG, Popejoy M, Evgeniev N, et al. (2023a). Efficacy and Safety of Adintrevimab (ADG20) for the Treatment of High-Risk Ambulatory Patients With Mild or Moderate Coronavirus Disease 2019: Results From a Phase 2/3, Randomized, Placebo-Controlled Trial (STAMP) Conducted During Delta Predominance and Early Emergence of Omicron. Open Forum Infect Dis. 10(6):ofad279. doi:10.1093/ofid/ofad279

24. Ison MG, Weinstein DF, Dobryanska M, et al. (2023b). Prevention of COVID-19 following a single intramuscular administration of adintrevimab: results from a phase 2/3 randomized, double-blind, placebo-controlled trial (EVADE). Open Forum Infect Dis. 10(7):ofad314. doi:10.1093/ofid/ofad314

25. Kang SW, Kim JW, Kim JY, et al. (2023). Characteristics and risk factors of prolonged viable virus shedding in immunocompromised patients with COVID-19: a prospective cohort study. J Infect. 86(4):412–414. doi:10.1016/j.jinf.2023.01.024

26. Ku JH, Sy LS, Qian L, et al. (2023). Vaccine effectiveness of the mRNA-1273 3-dose primary series against COVID-19 in an immunocompromised population: A prospective observational cohort study. Vaccine. 41(24):3636–3646. doi:10.1016/j.vaccine.2023.04.075

27. Lin KJ, Turner KC, Rosario M, et al. (2024). Population Pharmacokinetics of Casirivimab and Imdevimab in Pediatric and Adult Non-Infected Individuals, Pediatric and Adult Ambulatory or Hospitalized Patients or Household Contacts of Patients Infected with SARS-COV-2. Pharm Res. 41(10):1933–1949. doi:10.1007/s11095-024-03764-5

28. Link-Gelles R, Chickery S, Webber A, et al. (2025). Interim Estimates of 2024-2025 COVID-19 Vaccine Effectiveness Among Adults Aged >/=18 Years - VISION and IVY Networks, September 2024-January 2025. MMWR Morb Mortal Wkly Rep. 74(6):73–82. doi:10.15585/mmwr.mm7406a1

29. Merck & Co. (2024). Fact Sheet for Healthcare Providers: Emergency Use Authorization for Lagevrio (Molnupiravir) Capsules. Revised June 2024. https://www.fda.gov/media/155054/download

30. Montgomery H, Hobbs FDR, Padilla F, et al. (2022). Efficacy and safety of intramuscular administration of tixagevimab-cilgavimab for early outpatient treatment of COVID-19 (TACKLE): a phase 3, randomised, double-blind, placebo-controlled trial. Lancet Respir Med. 10(10):985–996. doi:10.1016/s2213-2600(22)00180-1

31. Pfizer. (2025). Fact Sheet for Health Care Providers: Emergency Use Authorization for Paxlovid. Updated February 2025. https://labeling.pfizer.com/ShowLabeling.aspx?id=16474&format=pdf

32. Regeneron. (2022). Fact Sheet for Health Care Providers: Emergency Use Authorization (EUA) of REGEN-COV (casirivimab and imdevimab). Updated January 2022. https://www.fda.gov/media/145611/download

33. Sager JE, El-Zailik A, Passarell J, et al. (2023). Population pharmacokinetics and exposure-response analysis of a single dose of sotrovimab in the early treatment of patients with mild to moderate COVID-19. CPT Pharmacometrics Syst Pharmacol. 12(6):853–864. doi:10.1002/psp4.12958

34. Schmidt P, Li Y, Popejoy M. (2024). Immunobridging for Pemivibart, a Monoclonal Antibody for Prevention of Covid-19. N Engl J Med. 391(19):1860–1862. doi:10.1056/NEJMc2404555

35. Stadler E, Chai KL, Schlub TE, et al. (2023). Determinants of passive antibody efficacy in SARS-CoV-2 infection: a systematic review and meta-analysis. Lancet Microbe. 4(11):e883–e892. doi:10.1016/s2666-5247(23)00194-5

36. Wolfe CR, Cohen J, Mahoney K, et al. (2025). Safety and Efficacy of Pemivibart, a Long-Acting Monoclonal Antibody, for Prevention of Symptomatic COVID-19: Interim Results From the CANOPY Clinical Trial Clinical Infectious Diseases. 10.1093/cid/ciaf265

